# Neurophobia: A study of Australian medical students and junior doctors

**DOI:** 10.1101/2021.02.21.21252144

**Authors:** Alex Yeung, Leila Karimi, Tissa Wijeratne

**Affiliations:** Department of Neurology, Australian Institute for Musculoskeletal Science, Level Three, Western Centre for Health Research and Education, Sunshine Hospital, Western Health & University Melbourne, St Albans, VIC, Australia; School of Psychology and Public Health, La Trobe University, Melbourne, VIC, Australia; Faculty of Social and Political Sciences, Ivane Javakhishvili Tbilisi State University, Tbilisi, Georgia; Department of Medicine, Faculty of Medicine, University of Rajarata, Anuradhapura, Sri Lanka

**Author notes:** **Corresponding Author:** Tissa Wijeratne Email. **Funding Source:** None. **Conflict of Interest Disclosure:** None.

**Keywords:** Neurology, Education, Neurophobia

## Abstract

**Background:** Neurological disorders are the leading cause disability in Australia and the world. Combating the perceived difficulty of neurology or “neurophobia” and improving physician education is a key component in addressing this problem. We aim to conduct the first study to identify whether neurophobia exists in medical students and junior doctors in an Australian population and try to identify factors that may contribute to this in this population.

**Methods:** A 24 question online validated survey was distributed via email broadcast to all medical students and junior doctors at a metropolitan tertiary care centre in Australia. Responses were collected over 6 weeks with weekly reminder emails for 4 weeks after the initial invitation email.

**Results:** 114 medical students and junior doctors participated in the study. Participants perceived neurology as the most difficult medical speciality compared to 10 other medical specialties (p=0.001). The top three factors contributing to this perceived difficulty were: a lack of understanding of neuroanatomy, lack of diagnostic certainty and lack of clinical exposure. 65% of the participants stated that they had too little planned teaching in neurology with only 36% of the participants having performed a neurology rotation during medical school.

**Conclusion:** The prevalence of neurophobia in this Australian cohort of medical students and junior doctors is consistent with previous findings from around the world. This concerning finding requires further examination into the contributing factors in order to created trials of targeted interventions in order to resolve this.

## Introduction

Neurological disorders (ND) are the leading cause of disability in the world[1]. NDs are the leading cause of disability in Australia as well. The historic 73^rd^ World Health Assembly (WHA) makes the strongest case for synergistic approach to tackle the NDs as a very high priority with 13 new paragraphs entailing the process further[2].

One in three of us are destined to end up with a stroke or dementia according the Framingham study[3]. This was elaborated at the recent WHA with the hope of prevention of NDs such as stroke, dementia, head injuries through proper education and intervention[2, 4, 5].

Physician education (specialist physicians as well as primary care physicians) in neurology is a key component of building synergy to address the global burden of neurological disorders. The concept of the “fear of clinical neurology” is a challenge in this regard.

The term “neurophobia”, or the concept of the fear of neuroscience and clinical neurology in medical students, was first published in the literature by Josefowicz in JAMA in 1994[6]. Since then the phenomenon of neurophobia has been found to be a common and prevailing theme in not only medical students but also in doctors (primary care as well as specialist care, across the board) throughout the world [7-10]. Published studies have not only demonstrated the high prevalence of neurophobia, but also investigated the factors that could be contributing to it and also possible educational interventions to try overcome neurophobia[7-12]. The current study is the first in the literature to the best of our knowledge and aims to identify whether neurophobia exists in medical students and junior doctors in an Australian population and tries to identify factors that may contribute to this in Australia.

## Methods

An online survey with 24 questions was used (Additional file 1), which was modelled closely on the validated survey used by Pakpoor[8]. This model was used to ensure comparability and consistency with previous studies investigating neurophobia. The survey consisted of a combination of multiple choice, short answer questions and Likert scale type questions. There were six basic demographic questions identifying the age, gender and details of the medical degree undertaken by the participant. This was followed by four questions used to identify neurophobia by comparing neurology with ten other medical specialties in terms of perceived difficulty in learning, clinical examination, drawing up a differential diagnosis, and rating of teaching quality. There were then fourteen questions enquiring about the participants’ perceptions of different aspects of their neurology education (including pre-clinical, clinical and research opportunities, and identification of areas of improvement), factors associated with the perceived difficulty of neurology, personal experience with neurological patients, and neurology as a career choice. Ethics approval for the study was provided by Western Health Ethics Committee, reference HREC/20/WH/61878. Western Health Neurophobia Survey (WHNS) was conducted according to the Helsinki declaration.

Surveys were distributed via email broadcast to all medical students and junior doctors (interns, basic physician trainees) at a metropolitan tertiary care centre in Australia. The invitation email included the rationale for the study, participant information and consent form (PICF) and link to the online survey via SurveyMonkey. Consent was implied via participation in the survey and all participation information was anonymous apart from the basic demographic information asked in the survey.

The survey was distributed in May 2020 with weekly reminder emails sent for four weeks after the initial invitation email. Responses were collected over 6 weeks from the first invitation email.

Independent t-tests were used to assess the significant differences between attitudes and barriers of neurology disciplines compared to other specialties in terms of perceived difficulty in learning neurology, perceived ability in examining patients with neurological disorders, comfort in developing a differential diagnosis and the quality of teaching received. Descriptive statistics such as percentages and frequencies were used to identify the most important factors in perceived difficulties in Neurology compared to other specialties. Chi-square data analysis procedures were used to identify the most important factors contributing to choosing Neurology as a career in the future. Data were analysed using SPSS (v 25). Qualitative thematic analysis was used to make sense of open-end responses.

## Results

The total number of participants included in the analysis was 114, which made a participation rate of 44% (260 invite emails). Table 1 presents the demographic characteristics of the participants. There was a slight majority of junior doctors (53%) over medical students (47%) who participated in the study. The majority of the participants (93%) had studied or were studying a postgraduate medical degree. 80% of the participants attended the same medical school. 56% of the participants were females while 44% of the participants were males with an average age of 26 years.

**Table 1.** Demographic characteristics of the participants (n=114)

| Characteristics | N (%) |
| --- | --- |
| Medical degree: |  |
| Postgraduate | 106 (93) |
| Undergraduate | 8 (7) |
| Year of medical school: |  |
| Year 2-3 | 42 (36.8) |
| Year 4-5 | 12 (10.5) |
| Junior Drs | 60 (52.6) |
| Gender: |  |
| Female | 64 (56.1) |
| Male | 50 (43.9) |
| Age mean (SD) | 26.09 (3.32) |

Participants rating of eleven medical specialities is demonstrated in table 2 based on their perceived difficulty, comfort with examination, comfort with drawing up a differential diagnosis and quality of teaching received. Participants perceived neurology as the most difficult of the eleven medical specialties (p =0.001). They had the least comfort in drawing up a differential diagnosis for patients presenting with neurological symptoms (p =0.001) compared to other medical specialties apart from haematology and nephrology. Discomfort with the neurological examination was rated by the participants as average (M (Sd)=3 (1.06)) and significantly higher (p=0.001) than five other specialities (i.e. respiratory, gastroenterology, cardiology, geriatrics and endocrinology). Participants’ rating of teaching quality for neurology was above the average and higher than 50% of other medical specialities (i.e. higher than haematology, rheumatology, infectious disease, oncology, nephrology). There were no significant differences between medical students and junior doctors in the above categories.

**Table 2.**
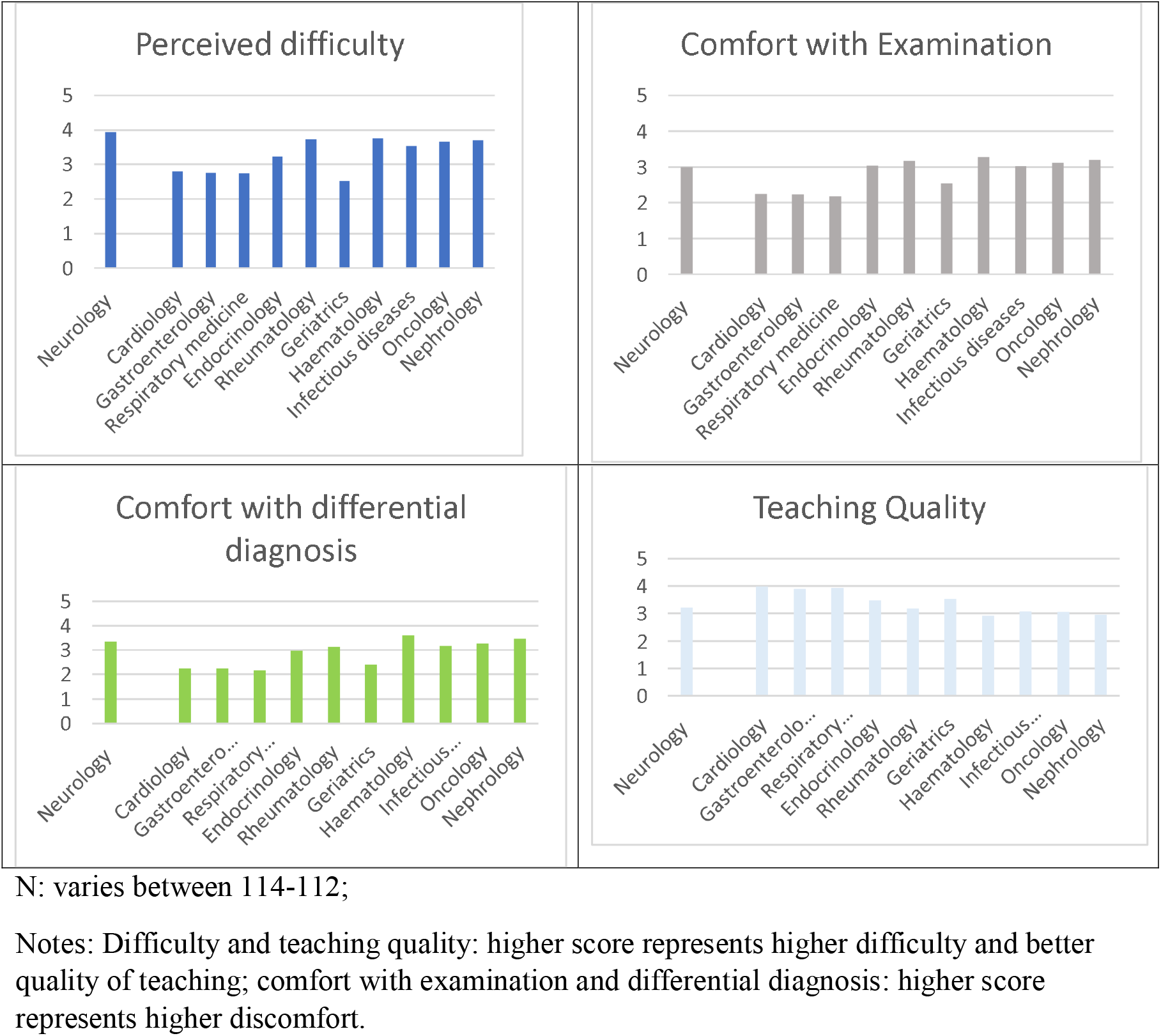
Comparing the participants’ ratings of perceived difficulty, comfort with examination and drawing up a differential diagnosis and the quality of teaching for eleven medical specialties.

The top three factors contributing to perceived difficulties of neurology were identified as a lack of understanding of neuroanatomy, lack of diagnostic certainty and lack of clinical exposure (figure 1). The factors were quite similar when compared between medical students and junior doctors except for the clinical neurological examination where it was perceived as more difficult for junior doctors compared to the students (p < 0.05).

**Figure 1.**
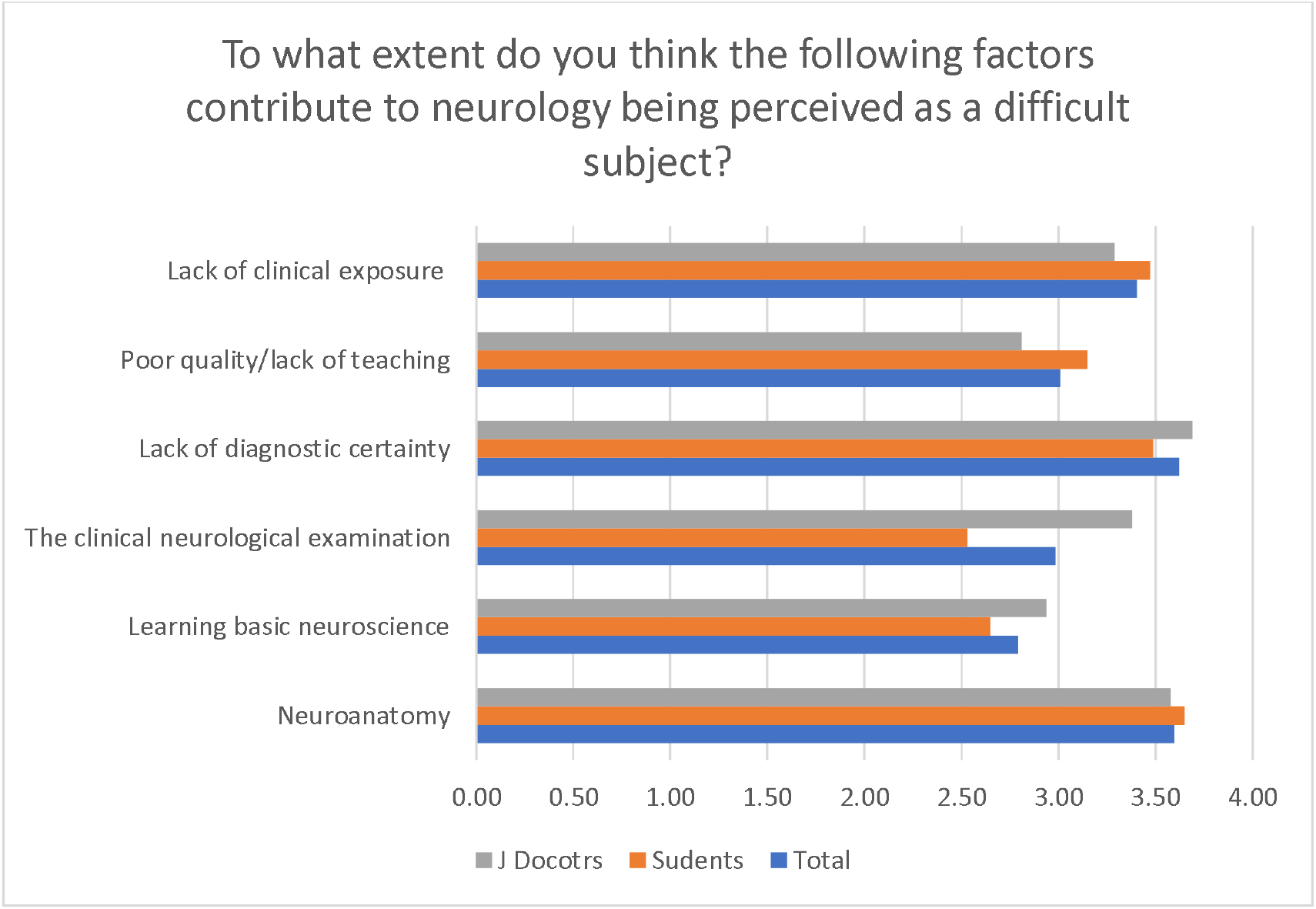
The contributing factors to neurology being perceived as a difficult subject (all participants, students vs. junior doctors)

In terms of neurology education, 65% of the participants stated they had had too little planned teaching in neurology. Outside of planned teaching, a significant proportion of participants stated they had not had an opportunity to receive additional neurology teaching beyond course curriculum (64%) or carry out a research project in a topic related to neurology (49%). Interestingly, only 36% of participants had performed a neurology rotation during medical school. In terms of improving medical education, the three options that received the highest ranking were: more/improved bedside teaching (38%), followed by more/improved textbooks (11.5%) and more/improved peer discussions (9%).

The top three contributing factors to choosing a career in neurology were research opportunity, the opportunity for career progression, and team-work followed closely by the ability to make a significant difference to patients’ lives (figure 2). Students significantly rated financial rewards, career progression and prestige higher than junior doctors (P<0.05). The most revealing finding was on the likelihood of pursuing a career in neurology which was rated as unlikely (M (sd)=2.09 (.81)).

## Discussion

This study is first to identify the existence of neurophobia in an Australian cohort of medical students and junior doctors. Similar to previous studies, neurology was perceived as the most difficult medical specialty with higher levels of discomfort with drawing up a differential diagnosis compared to other specialties[7, 8]. This was despite perceived adequate comfort with the neurological examination and teaching quality.

In terms of the factors which contributed to neurology’s perceived difficulties, two of the top three factors were identical to those reported by in Pakpoor’s study (neuroanatomy, lack of diagnostic certainty)[8], with neuroanatomy being the biggest factor followed by lack of diagnostic certainty and lack of clinical exposure. There were also eight qualitative answers for this question, of which half of them commented on the complexity of reaching a diagnosis due to various factors, e.g. perceived subjectiveness of taking the history and performing the examination and integrating these clinical findings with the wide array of neurological conditions.

Interestingly, despite rating adequate teaching quality, the majority of participants stated that they had had too little planned teaching in neurology and did not have opportunities for further teaching or research outside of the medical school curriculum. One of the most striking statistics was that only 36% of participants had stated that they had performed a neurology rotation during medical school. This was an unexpectedly low percentage even after accounting for a proportion of the participants who had not yet commenced clinical rotations (16%).

In terms of improving neurology education, they identified that more/improved bedside teaching was by far the most popular suggestion. One qualitative answer was provided to this topic which stated that more dedicated clinical teaching during clinical years of medical school would be more useful.

In terms of the general qualitative comments that were able to be provided by the participants, there was a large heterogeneity of content that was brought up, but the two most common themes included: the complexity of neurological conditions (including their neuroanatomical and pathophysiological bases) and the lack of clinical exposure/teaching (including neurology rotations and bedside tutorials). It is possible that a combination of these factors led not only to the perception of high difficulty of neurology but also the low likelihood of participants to pursue a career in neurology.

The main limitations of this study include: the small sample size and homogeneity of this cohort as the questionnaire was only distributed to medical students and junior doctors from a single centre of which 80% of the participants were undergoing/had undergone their medical training through one medical school. As a result, these results may not be generalisable to similar cohorts throughout Australia. Secondly, the participation rate of 44% (although typical for these studies) meant that we were not able to collect data from the majority of the cohort which leads the results to be subject to possible biases, predominantly selection bias, as the participants may have been more likely to participate if they did suffer from neurophobia due to their identification with the study topic.

Synthesising the data collected from this study, we appreciate that possible targeted interventions in combating neurophobia in this cohort would be a change to neurology education during clinical years including mandatory clinical neurology rotations and structured clinical/bedside tutorials. Drawing from the successes of Ridsdale[12], it would not be unreasonable to suggest that the education in clinical years should be taught by neurologists who would be able to simplify the perceived “complexity” of neurology due to their greater understanding of the subject matter.

## Conclusion

This study is first to identify the existence of neurophobia in an Australian cohort of medical students and junior doctors. Various contributing factors identified by the participants were similar to those identified in other cohorts whom also suffered from neurophobia. It is worth noting the fact that approximately two thirds of our future doctors are not exposed to a clinical neurology rotation throughout their training despite access to a busy, neurology service attached to the Victorian public hospital (well over 6,000 neurology and stroke inpatient visits per year with eleven subspecialty clinics covering all aspects of neurology with immense learning opportunity). This is despite the fact that NDs are the leading cause of disability in Australia as shown by the Global Burden of Diseases publications year after year. Further studies in this population are required to explore these factors and create targeted educational interventions to combat this. Our study makes a strong case for the value of exposing medical students to clinical neurology rotations and research opportunities as a matter of priority.

## Data Availability

Data will be available through the corresponding author with a reasonable request.

## Abbreviations

(ND): Neurological disorders
(WHA): World Health Assembly
JAMA: (The Journal of the American Medical Association)
(WHNS): Western Health Neurophobia Survey
(PICF): participant information and consent form

## Acknowledgements

None

